# RNA expression data suggest ALDH2 inhibition as a concept for killing acute myeloid leukemia cells that are resistant to ALDH1 inhibitors: the example of NPM1 mutant AML

**DOI:** 10.1101/2024.11.09.24317037

**Authors:** Garrett M Dancik, Spiros Vlahopoulos

**Affiliations:** Department of Computer Science, Eastern Connecticut State University, Willimantic, CT 06226, USA; First Department of Pediatrics, National and Kapodistrian University of Athens, Thivon & Levadeias 8, Goudi, 11527 Athens, Greece

## Abstract

In this data paper we present a study of RNA expression in association with the disease course of acute myeloid leukemia (AML). We have previously identified aldehyde dehydrogenase genes *ALDH1A1* and *ALDH2* as prospective actionable targets in AML. ALDH1A1 is expected to have key functions in leukemia stem-like cells that are prone to a dormant state in terms of metabolic activity and proliferation. Cells with a higher activity of metabolism as a whole and in mitochondria in particular, are likely to generate a higher abundance of formaldehyde and acetaldehyde. Cell survival necessitates removal of formaldehyde and acetaldehyde, which is to a substantial degree the function of ALDH2. AML cells with a mutant *NPM1* gene permit higher MYC activity which would lead to increased metabolic activity. Extended MYC activity is allowed by the mutant NPM1 protein, compared to NPM1 wild-type. Here, we show that survival analysis in patients with *NPM1* mutant AML yields a higher hazard ratio for *ALDH2* RNA than for *ALDH1A1*, which is not the case for patients with wild-type NPM1. This result is consistent with the difference in enzymatic function between ALDH2 and ALDH1A1, the latter of which is not suited for small aldehydes, especially formaldehyde. This should open the door to the examination of ALDH2 inhibitors such as the clinically approved disulfiram, for the treatment of NPM1 mutant AML that proves refractory to ALDH1A1 inhibition.

## Main text

We have previously suggested that the aldehyde dehydrogenase protein family, and especially members ALDH1A1 and ALDH2, constitute actionable targets for translational research in acute myeloid leukemia (AML), because the respective genes *ALDH1A1* and *ALDH2* encode RNA which are frequently overexpressed in patients with poor prognosis [1]. Very importantly, others have shown that a) patients with CD34(+)CD38(-) leukemia cells with high aldehyde dehydrogenase activity manifest a significantly lower complete remission rate, as well as poorer event-free and overall survival [2] and b) importantly, ALDH1 inhibition with agent DIMATE kills preferably AML stem cells, without comparable toxicity for normal hematopoietic cells [3]. Furthermore this agent (DIMATE) is currently studied in a Phase I clinical trial for AML [4].

Based on the literature, we previously postulated that in respect to metabolism and oxidative stress, there are at least two basic phenotypic states for leukemia stem-like cells; these extreme states present variable degrees of overlap in a given cell or in daughter clones [5] :

1) a state of relative metabolic dormancy with varying degrees of cycling quiescence and a protection from oxidant stress mainly from a decreased generation of reactive oxygen species, and secondarily from increased expression and activity of cytosolic retinaldehyde enzymes such as ALDH1A1. It must be noted that quiescence can be induced in leukemia cells or in naive embryonic stem cells by inhibition of Myc, which among several other effects is accompanied by decreased one-carbon metabolism, and decreased oxidant stress [6] [7] [8] [9].

2) a state of increased metabolic activity and high capacity for proliferation, characterized by high activity of transcription factors such as Myc, and a protection from oxidant stress mainly through a number of potent antioxidant enzymatic systems. A Myc-dominated cellular phenotype should generate large amounts of formaldehyde and acetaldehyde through several metabolic processes [10] [11] [12] [13] [14] [15]. Formaldehyde and acetaldehyde in mitochondria can be removed by ALDH2 [16] [14].

The turnover of Myc is controlled also by Nucleophosmin, encoded by gene *NPM1*, which is frequently mutated in AML, extending Myc activity [17] [18]. Among a number of advanced methods to interfere with the effects of *NPM1* mutation, one method is via intervention targeting the scaffold protein menin [19]. It was shown that mutated NPM1 protein, remaining in the cytoplasm, enhances leukemia progression via permitting induction of *HOX* gene expression [20]. In fact, nuclear relocalization or degradation of mutant NPM1 induces cell growth arrest and differentiation in primary AML cells. It was shown that NPM1 mutant protein conferred to AML cells a growth advantage [20].

The data we present here suggest that there a feasible and additional approach to targeting AML cells with increased metabolic activity and proliferation through inhibition of the mitochondrial enzyme ALDH2. This inhibition can be expected to affect those malignant cells that rely on mitochondrial activity for energy generation, and thus generate formaldehyde in substantial abundance [11]. In zebrafish cells formaldehyde accumulation was shown to cause organelle damage [21].

Here we examine data from published microarray-based studies of AML patient samples, specifically the Gene Expression Omnibus (public functional genomics data repository supporting MIAME-compliant data submissions) Datasets GSE 1159, 242, 561, 536, and 34 with the help of the online analysis platform KM Plotter [22] (https://kmplot.com/analysis/index.php?p=background; accessed on 30 October 2024), and find that stratification of patients based on *ALDH2* RNA expression yields a higher hazard ratio than stratification based on *ALDH1A1* RNA expression, for both overall, event-free, and post-progression free survival in *NPM1* mutant AML (Table 1). Furthermore, the hazard ratio for *ALDH2* RNA expression is higher in *NPM1* mutant patients compared to wild-type, which is not the case with *ALDH1A1*.

**Table 1.** Stratification of patients based on *ALDH1A1* and *ALDH2* RNA expression.

|  |  |  |  |
| --- | --- | --- | --- |
| **OVERALL SURVIVAL** | AML TOTAL (HR, P, FDR) | NMP1 WT (HR, P, FDR) | NPM1MT (HR, P, FDR) |
| *ALDH1A1* | 1.47, 1.3e-10, 1% | 1.99, 6.6e-10, 1% | 1.21, 0.3, 100% |
| *ALDH2* | 1.81, <1e-16, 1% | 2.08, 4.4e-9, 1% | 2.19, 0.00012, 1% |
| **EVENT FREE SURVIVAL** | AML TOTAL (HR, P, FDR) | NMP1 WT (HR, P, FDR) | NPM1MT (HR, P, FDR) |
| *ALDH1A1* | 1.7, 2.1e-6, 1% | 2.43, 1.8e-9, 1% | 0.79, 0.26, 100% |
| *ALDH2* | 1.75, 6.1e-5, 1% | 1.98, 1.1e-5, 1% | 2.35, 0.00029, 2% |
| **POST PROGRESSION SURVIVAL** | AML TOTAL (HR, P, FDR) | NMP1 WT (HR, P, FDR) | NPM1MT (HR, P, FDR) |
| *ALDH1A1* | 1.61, 0.00063, 5% | 1.54, 0.0083, >50% | 1.53, 0.097, 100% |
| *ALDH2* | 1.43, 0.0056, 50% | 1.54, 0.016, >50% | 1.95, 0.0031, 10% |

Abbreviations: HR, hazard ratio; FDR, false discovery rate

What makes this result even more compelling is the fact that when we look individually at the datasets with known *NPM1* mutants, and stratify patients based on *ALDH1A1* or *ALDH2* expression, we note that the difference between *NPM1* wild-type and mutant AML generally holds true. In both datasets, stratification of patients with NPM1 mutations based on *ALDH2* RNA expression yields a higher hazard ratio than stratification based on *ALDH1A1* RNA expression, for both overall and event-free survival. (Table 2).

**Table 2.** Stratification of patients from the studies GSE1159 and GSE6891, based on *ALDH1A1* and *ALDH2* RNA expression.

| **OVERALL SURVIVAL** | AML TOTAL (HR, P, FDR) | NMP1 WT (HR, P, FDR) | NPM1MT (HR, P, FDR) |
| --- | --- | --- | --- |
| **GSE1159** |  |  |  |
| *ALDH1A1* | 1.62, 0.0014, 20% | 2.37, 3.7e-6, 1% | 1.25, 0.46, 100% |
| *ALDH2* | 2.08, 4.4e-9, 1% | 2.31, 8.3e-6, 1% | 1.94, 0.024, >50% |
| **GSE6891** |  |  |  |
| *ALDH1A1* | 1.51, 0.00013, 1% | 1.92, 2.3e-6, 1% | 1.27, 0.28, 100% |
| *ALDH2* | 1.92, 4.3e-8, 1% | 2.06, 7e-6, 1% | 2.62, 0.00069, 5% |
| **EVENT FREE SURVIVAL** | AML TOTAL (HR, P, FDR) | NMP1 WT (HR, P, FDR) | NPM1MT (HR, P, FDR) |
| **GSE1159** |  |  |  |
| *ALDH1A1* | 1.6, 0.021, >50% | 2.4, 5e-4, 3% | 0.63, 0.18, 100% |
| *ALDH2* | 1.9, 0.0021, 20% | 2.49, 0.00039, 2% | 3.19, 0.022, 50% |
| **GSE6891** |  |  |  |
| *ALDH1A1* | 1.82, 9.9e-6, 1% | 2.47, 5.9e-7, 1% | 0.71, 0.2, 100% |
| *ALDH2* | 1.6, 0.00098, 10% | 1.84, 0.0014, 10% | 2.84, 0.0016, 10% |
| **POST PROGRESSION SURVIVAL** | AML TOTAL (HR, P, FDR) | NMP1 WT (HR, P, FDR) | NPM1MT (HR, P, FDR) |
| **GSE1159** |  |  |  |
| *ALDH1A1* | 1.53, 0.05, >50% | 1.79, 0.04, >50% | 2.05, 0.14, 100% |
| *ALDH2* | 1.49, 0.09, 100% | 2.12, 0.0087, 20% | 1.82, 0.22, 100% |
| **GSE6891** |  |  |  |
| *ALDH1A1* | 1.7, 0.0012, 10% | 1.71, 0.0079, 50% | 1.48, 0.2, 100% |
| *ALDH2* | 1.5, 0.014, >50% | 1.69, 0.029, >50% | 2.23, 0.0048, 10% |

Abbreviations: HR, hazard ratio; FDR, false discovery rate

The most common clinically approved inhibitor for ALDH2 is disulfiram (used for the maintenance of abstinence from alcohol), which should cause accumulation of toxic formaldehyde in cells that operate mitochondrial respiratory chain with high activity, and is shown to cause accumulation of acetaldehyde in cells, in general [23] [24] [25] [26] [27]. This would make disulfiram an important translational agent for AML cells resistant to ALDH1 (retinaldehyde dehydrogenase) inhibitors because ALDH1 inhibitors would tend to have a stronger effect against more quiescent leukemia cells. This could have a broader significance due to a number of alterations that bestow cancer cells with the capacity to respond to favorable changes in conditions with a rapid increase in metabolic capacity and macromolecule turnover and repair [28] [29] [30] [31].

We previously discovered that from the nineteen members of the ALDH family, ALDH1A1 and ALDH2 have the strongest association with AML patient risk group classification [1]. Our current findings support the hypothesis that ALDH1A1 is the gene product most likely to protect quiescent stem-like AML cells, while ALDH2 is the gene product most likely to protect proliferating AML cells, regardless of whether these cells manifest stem-like properties or not. A probable reason for the importance of ALDH2 in leukemia is the detoxification of small aldehydes, especially formaldehyde.

## Data Availability

All data produced are available online from https://kmplot.com/analysis/index.php?p=service&cancer=aml.

## Notes

### Competing Interest Statement

The authors have declared no competing interest.

### Funding Statement

This study did not receive any funding

## References

1. Dancik, G.M.; Varisli, L.; Tolan, V.; Vlahopoulos, S. Aldehyde Dehydrogenase Genes as Prospective Actionable Targets in Acute Myeloid Leukemia. Genes 2023, 14, 1807, doi:10.3390/genes14091807.

2. Gerber, J.M.; Zeidner, J.F.; Morse, S.; Blackford, A.L.; Perkins, B.; Yanagisawa, B.; Zhang, H.; Morsberger, L.; Karp, J.; Ning, Y.; et al. Association of Acute Myeloid Leukemia’s Most Immature Phenotype with Risk Groups and Outcomes. Haematologica 2016, 101, 607–616, doi:10.3324/haematol.2015.135194.

3. Venton, G.; Pérez-Alea, M.; Baier, C.; Fournet, G.; Quash, G.; Labiad, Y.; Martin, G.; Sanderson, F.; Poullin, P.; Suchon, P.; et al. Aldehyde Dehydrogenases Inhibition Eradicates Leukemia Stem Cells While Sparing Normal Progenitors. Blood Cancer J. 2016, 6, e469, doi:10.1038/bcj.2016.78.

4. Venton, G.; Colle, J.; Tichadou, A.; Quessada, J.; Baier, C.; Labiad, Y.; Perez, M.; De Lassus, L.; Loosveld, M.; Arnoux, I.; et al. Reactive Oxygen Species and Aldehyde Dehydrogenase 1A as Prognosis and Theragnostic Biomarker in Acute Myeloid Leukaemia Patients. J. Cell. Mol. Med. 2024, 28, e70011, doi:10.1111/jcmm.70011.

5. Vlahopoulos, S.; Pan, L.; Varisli, L.; Dancik, G.M.; Karantanos, T.; Boldogh, I. OGG1 as an Epigenetic Reader Affects NFκB: What This Means for Cancer. Cancers 2023, 16, 148, doi:10.3390/cancers16010148.

6. Takeishi, S.; Matsumoto, A.; Onoyama, I.; Naka, K.; Hirao, A.; Nakayama, K.I. Ablation of Fbxw7 Eliminates Leukemia-Initiating Cells by Preventing Quiescence. Cancer Cell 2013, 23, 347–361, doi:10.1016/j.ccr.2013.01.026.

7. Aleksandrova, K.V.; Vorobev, M.L.; Suvorova, I.I. mTOR Pathway Occupies a Central Role in the Emergence of Latent Cancer Cells. Cell Death Dis. 2024, 15, 176, doi:10.1038/s41419-024-06547-3.

8. Chen, C.; Liu, Q.; Chen, W.; Gong, Z.; Kang, B.; Sui, M.; Huang, L.; Wang, Y.-J. PRODH Safeguards Human Naive Pluripotency by Limiting Mitochondrial Oxidative Phosphorylation and Reactive Oxygen Species Production. EMBO Rep. 2024, 25, 2015–2044, doi:10.1038/s44319-024-00110-z.

9. Khoa, L.T.P.; Yang, W.; Shan, M.; Zhang, L.; Mao, F.; Zhou, B.; Li, Q.; Malcore, R.; Harris, C.; Zhao, L.; et al. Quiescence Enables Unrestricted Cell Fate in Naive Embryonic Stem Cells. Nat. Commun. 2024, 15, 1721, doi:10.1038/s41467-024-46121-1.

10. Purhonen, J.; Klefström, J.; Kallijärvi, J. MYC-an Emerging Player in Mitochondrial Diseases. Front. Cell Dev. Biol. 2023, 11, 1257651, doi:10.3389/fcell.2023.1257651.

11. Tenney, L.; Pham, V.N.; Brewer, T.F.; Chang, C.J. A Mitochondrial-Targeted Activity-Based Sensing Probe for Ratiometric Imaging of Formaldehyde Reveals Key Regulators of the Mitochondrial One-Carbon Pool. Chem. Sci. 2024, 15, 8080–8088, doi:10.1039/d4sc01183j.

12. Nikiforov, M.A.; Chandriani, S.; O’Connell, B.; Petrenko, O.; Kotenko, I.; Beavis, A.; Sedivy, J.M.; Cole, M.D. A Functional Screen for Myc-Responsive Genes Reveals Serine Hydroxymethyltransferase, a Major Source of the One-Carbon Unit for Cell Metabolism. Mol. Cell. Biol. 2002, 22, 5793–5800, doi:10.1128/MCB.22.16.5793-5800.2002.

13. Antoshechkin, A.G. Physiological Model of the Stimulative Effects of Alcohol in Low-to-Moderate Doses. Ann. N. Y. Acad. Sci. 2002, 957, 288–291, doi:10.1111/j.1749-6632.2002.tb02927.x.

14. Dancik, G.M.; Voutsas, I.F.; Vlahopoulos, S. Aldehyde Dehydrogenase Enzyme Functions in Acute Leukemia Stem Cells. Front. Biosci. Sch. Ed. 2022, 14, 8, doi:10.31083/j.fbs1401008.

15. Arumugam, M.K.; Gopal, T.; Kalari Kandy, R.R.; Boopathy, L.K.; Perumal, S.K.; Ganesan, M.; Rasineni, K.; Donohue, T.M.; Osna, N.A.; Kharbanda, K.K. Mitochondrial Dysfunction-Associated Mechanisms in the Development of Chronic Liver Diseases. Biology 2023, 12, 1311, doi:10.3390/biology12101311.

16. Klyosov, A.A. Kinetics and Specificity of Human Liver Aldehyde Dehydrogenases toward Aliphatic, Aromatic, and Fused Polycyclic Aldehydes. Biochemistry 1996, 35, 4457–4467, doi:10.1021/bi9521102.

17. Bonetti, P.; Davoli, T.; Sironi, C.; Amati, B.; Pelicci, P.G.; Colombo, E. Nucleophosmin and Its AML-Associated Mutant Regulate c-Myc Turnover through Fbw7 Gamma. J. Cell Biol. 2008, 182, 19–26, doi:10.1083/jcb.200711040.

18. Lai, Q.; Hamamoto, K.; Luo, H.; Zaroogian, Z.; Zhou, C.; Lesperance, J.; Zha, J.; Qiu, Y.; Guryanova, O.A.; Huang, S.; et al. NPM1 Mutation Reprograms Leukemic Transcription Network via Reshaping TAD Topology. Leukemia 2023, 37, 1732–1736, doi:10.1038/s41375-023-01942-9.

19. Patel, S.S. NPM1-Mutated Acute Myeloid Leukemia: Recent Developments and Open Questions. Pathobiol. J. Immunopathol. Mol. Cell. Biol. 2024, 91, 18–29, doi:10.1159/000530253.

20. L, B.; Mc, G.; D, S.; Ag, G.; Yh, H.; R, R.; I, G.; F, M.; F, M.; B, N.; et al. Mutant NPM1 Maintains the Leukemic State through HOX Expression. Cancer Cell 2018, 34, doi:10.1016/j.ccell.2018.08.005.

21. Xin, F.; Tian, Y.; Gao, C.; Guo, B.; Wu, Y.; Zhao, J.; Jing, J.; Zhang, X. A Two-Photon Fluorescent Probe for Basal Formaldehyde Imaging in Zebrafish and Visualization of Mitochondrial Damage Induced by FA Stress. The Analyst 2019, 144, 2297–2303, doi:10.1039/c8an02108b.

22. Győrffy, B. Discovery and Ranking of the Most Robust Prognostic Biomarkers in Serous Ovarian Cancer. GeroScience 2023, 45, 1889–1898, doi:10.1007/s11357-023-00742-4.

23. Human Endogenous Formaldehyde as an Anticancer Metabolite: Its Oxidation Downregulation May Be a Means of Improving Therapy - PubMed Available online: https://pubmed.ncbi.nlm.nih.gov/30370669/ (accessed on 6 November 2024).

24. Komarova, T.V.; Sheshukova, E.V.; Kosobokova, E.N.; Kosorukov, V.S.; Shindyapina, A.V.; Lipskerov, F.A.; Shpudeiko, P.S.; Byalik, T.E.; Dorokhov, Y.L. The Biological Activity of Bispecific Trastuzumab/Pertuzumab Plant Biosimilars May Be Drastically Boosted by Disulfiram Increasing Formaldehyde Accumulation in Cancer Cells. Sci. Rep. 2019, 9, 16168, doi:10.1038/s41598-019-52507-9.

25. Umansky, C.; Morellato, A.E.; Rieckher, M.; Scheidegger, M.A.; Martinefski, M.R.; Fernández, G.A.; Pak, O.; Kolesnikova, K.; Reingruber, H.; Bollini, M.; et al. Endogenous Formaldehyde Scavenges Cellular Glutathione Resulting in Redox Disruption and Cytotoxicity. Nat. Commun. 2022, 13, 745, doi:10.1038/s41467-022-28242-7.

26. Tang, C.; Pang, X.; Guo, Z.; Guo, R.; Liu, L.; Chen, X. Dual Action of Acidic Microenvironment on the Enrichment of the Active Metabolite of Disulfiram in Tumor Tissues. Drug Metab. Dispos. Biol. Fate Chem. 2021, 49, 434–441, doi:10.1124/dmd.120.000317.

27. Zhu, L.; Pei, W.; Thiele, I.; Mahadevan, R. Integration of a Physiologically-Based Pharmacokinetic Model with a Whole-Body, Organ-Resolved Genome-Scale Model for Characterization of Ethanol and Acetaldehyde Metabolism. PLoS Comput. Biol. 2021, 17, e1009110, doi:10.1371/journal.pcbi.1009110.

28. Reikvam, H. Inhibition of NF-κB Signaling Alters Acute Myelogenous Leukemia Cell Transcriptomics. Cells 2020, 9, 1677, doi:10.3390/cells9071677.

29. Rae, C.; Langa, S.; Tucker, S.J.; MacEwan, D.J. Elevated NF-kappaB Responses and FLIP Levels in Leukemic but Not Normal Lymphocytes: Reduction by Salicylate Allows TNF-Induced Apoptosis. Proc. Natl. Acad. Sci. U. S. A. 2007, 104, 12790–12795, doi:10.1073/pnas.0701437104.

30. Vlahopoulos, S.A. Divergent Processing of Cell Stress Signals as the Basis of Cancer Progression: Licensing NFκB on Chromatin. Int. J. Mol. Sci. 2024, 25, 8621, doi:10.3390/ijms25168621.

31. Ke, B.; Li, A.; Fu, H.; Kong, C.; Liu, T.; Zhu, Q.; Zhang, Y.; Zhang, Z.; Chen, C.; Jin, C. PARP-1 Inhibitors Enhance the Chemosensitivity of Leukemia Cells by Attenuating NF-кB Pathway Activity and DNA Damage Response Induced by Idarubicin. Acta Biochim. Biophys. Sin. 2022, 54, 91–98, doi:10.3724/abbs.2021011.

